# Osteoarthritis Diagnoses are Under-recorded in Primary Care Electronic Health Records in New Zealand

**DOI:** 10.1101/2025.11.20.25340701

**Authors:** Ross Wilson, Rory McMahon Chistopherson, Yana Pryymachenko, Richelle Caya, Lívia Gaspar Fernandes, Yen Wei Lim, Moody Gayed, Andrés Pierobon, Tony Dowell, Ben Darlow, Jayden MacRae, J. Haxby Abbott

## Abstract

**Background:** Effective planning of healthcare services for osteoarthritis requires accurate estimates of its prevalence in primary care. There is growing evidence to suggest that osteoarthritis prevalence estimates in primary care are inaccurate owing to under-recording of codified diagnoses within electronic health records.

**Aim:** To quantify the gap in recording of osteoarthritis diagnoses in primary care in New Zealand.

**Methods:** We linked data from the electronic health records of a cohort of 72,861 patients enrolled with primary care providers in the lower North Island of New Zealand with public hospital discharge records to identify all patients in the cohort with a total hip or knee replacement surgery for osteoarthritis between 2013-2018. We then calculated the proportion of patients who had at least one matching osteoarthritis diagnosis code in the primary care records prior to joint replacement surgery. We conducted subgroup analyses stratifying the cohort by joint, year of surgery, age, sex, ethnicity, socioeconomic deprivation, primary care practice, and frequency of contact with the practice.

**Results:** Among the 591 patients with hip replacement and 609 with knee replacement, 48.2% and 49.8% respectively had an osteoarthritis diagnosis code in the primary care records prior to the time of surgery. There was little difference between the rates of osteoarthritis diagnosis coding for demographic subgroups, but wide variations were observed amongst practices.

**Conclusions:** There is substantial under-recording of osteoarthritis diagnoses in primary care in New Zealand due to delayed or absent coding within electronic health records. Efforts should be made to improve recording of osteoarthritis diagnoses to aid in future policy decision-making and research.

## Introduction

Osteoarthritis (OA) is a leading global cause of disability associated with an array of personal costs, such as pain, impaired mobility, and reduced quality of life, along with various societal costs including increased healthcare spending and reduced labour force productivity^1–3^. Moreover, the burden of OA is projected to rise, casting further pressure upon healthcare systems to keep up with growing demand^1,4–6^.

Service delivery efforts to manage the growing burden of OA require astute planning, resource allocation decision-making, and health economic evaluation^7^, all of which rely on sound estimates of the prevalence of OA. As OA is generally diagnosed and managed in primary care^8^, reliable estimates of the prevalence and management of OA in primary care settings are crucial for effective service delivery planning. However, there is growing evidence to suggest existing data on the prevalence of OA in primary care are inaccurate^9–11^.

Diagnosis of OA is typically made on clinical grounds, without requiring confirmation by imaging^12–14^, and there are no pharmacological treatments specific to OA, so identification of patients in electronic health records (EHRs) relies on accurate diagnostic coding. International evidence has shown the coding of OA diagnoses within primary care EHRs to be inconsistent^9–11,5^, and there is preliminary evidence of under-recording of OA diagnoses in New Zealand (NZ) primary care settings^16^.

The aim of the present study is to quantify the extent to which OA diagnoses are under-recorded within primary care in NZ, by comparison with public hospital discharge records of patients who had a total hip or knee replacement surgery.

## Methods

### Data sources

This retrospective descriptive study used data held in the NZ Integrated Data Infrastructure (IDI). The IDI is a linked research database maintained by Stats NZ, NZ’s national statistics agency, containing individual-level data from government administrative datasets, Stats NZ surveys and data collections, and non-governmental organisations^17^.

The IDI contains, *inter alia*, extensive data from secondary care services (including public hospital discharge records), but primary care data are lacking. For this study, primary care EHRs pertaining to OA diagnosis were extracted from data held by Tū Ora Compass Health, a Primary Healthcare Organisation (PHO) in the lower North Island of NZ, and were integrated with the IDI (via record linkage at the individual patient level).

The cohort used in this study consisted of all patients who had total joint replacement (TJR) surgery of the hip or knee with a primary diagnosis of OA between January 2013 and December 2018, whose data were available in the extracted PHO data, and who were enrolled with any General Practitioner (GP) practice within Tū Ora Compass Health continuously for at least 10 years up to the date of surgery. Hip and knee replacement surgeries were treated as separate events, so an individual with both a hip and a knee replaced during the study period would contribute two independent observations. If a patient had more than one of the same procedure (e.g., both knees replaced in separate procedures), only the first eligible surgery was included.

### Variables

TJR surgeries were identified from hospital discharge records using the Australian Refined-Diagnosis Related Groups classification (AR-DRG, versions 6.0-7.0; codes I03A/B [hip replacement], I04A/B [knee replacement]), and associated OA diagnoses using the International Classification of Diseases and Related Health Problems, Australian Modification (ICD-10-AM; codes M16 [hip OA], M17 [knee OA]).

The outcome of interest was a coded diagnosis of OA of the same joint in the primary care EHR at any time prior to surgery. While all included patients were required to have at least 10 years of continuous PHO enrolment, the primary care data also included historical OA diagnosis codes recorded by a practice within the PHO at any time. OA diagnoses in primary care records were identified using Read codes (hip OA: 7K30*, 7K31*, 7K32*, 7K37*, 7K39*, N05z5.11, N05zJ.00; knee OA: 7K22*, N05zL.00, N05z6.11), supplemented as necessary with free-text annotations to differentiate hip and knee OA from OA in other body locations^18^.

### Data availability and ethics statement

The anonymised data used in this study were made available to the research team by Stats NZ under strict data access requirements and are not publicly available. The aggregated data and code used to produce the results reported in the article are available at https://osf.io/nfy5v/. For more information regarding access to the underlying raw data in the IDI, see https://www.stats.govt.nz/integrated-data/. To align with the confidentiality provisions for IDI data access, all reported counts of individuals are randomly rounded (up or down) to the nearest multiple of three.

The study was approved by the University of Otago Human Research Ethics Committee (Health) (reference HD18/077).

### Analyses

We calculated the proportion of OA TJR patients who had at least one matching prior OA diagnosis code in the primary care records. For additional exploratory analyses, we also calculated the time between first recorded diagnosis and the date of surgery, including for OA diagnoses occurring after surgery.

Subgroup analyses were conducted by stratifying the cohort by year of surgery, OA joint, age, sex, ethnicity (Māori, Pacific Islander, Asian), GP practice, socioeconomic deprivation (measured at the ‘meshblock’ level, representing small neighbourhoods of approximately 100-200 people), and frequency of contact with the GP practice.

All analyses were conducted using R version 4.4.0^19^.

## Results

The PHO cohort consisted of 72 861 patients, of whom 1593 had undergone TJR between 2013 and 2018. After excluding patients who were not enrolled with the PHO continuously for at least 10 years prior to surgery, the analysis cohort consisted of 1170 patients, with a total of 1 200 TJRs (591 hip replacements and 609 knee replacements). The mean (SD) age of the cohort was 69.5 (9.5) years and slightly more than half (57.5%) were female (Table 1).

**Table 1:** Demographic characteristics of the cohort.

| Variable | Value |
| --- | --- |
| N | 1200 |
| Joint |  |
| Hip | 591 (49.2%) |
| Knee | 609 (50.7%) |
| Age, mean (SD) | 69.5 (9.5) |
| Under 60 | 192 (16.0%) |
| 60 to 64 | 147 (12.2%) |
| 65 to 69 | 237 (19.8%) |
| 70 to 74 | 234 (19.5%) |
| 75 to 79 | 213 (17.8%) |
| 80 and over | 174 (14.5%) |
| Female | 690 (57.5%) |
| Ethnicity |  |
| Māori | 93 (7.8%) |
| Pacific Islander | 36 (3.0%) |
| Asian | 24 (2.0%) |
| NZ Deprivation Index, mean (SD) | 4.9 (2.6) |
| Decile 1 | 123 (10.2%) |
| Decile 2 | 147 (12.2%) |
| Decile 3 | 153 (12.8%) |
| Decile 4 | 150 (12.5%) |
| Decile 5 | 141 (11.8%) |
| Decile 6 | 114 (9.5%) |
| Decile 7 | 141 (11.8%) |
| Decile 8 | 117 (9.8%) |
| Decile 9 | 87 (7.2%) |
| Decile 10 | 27 (2.2%) |

Values are count (%) unless otherwise specified. All variables measured at the time of surgery. Abbreviations: NZ=New Zealand; SD=Standard deviation.

Of the 1 200 recorded procedures, 48.8% had a matching OA diagnosis code in the primary care EHR prior to the date of surgery. The rate was similar between hip (48.2%) and knee (49.8%) replacement surgeries. The rate of prior OA diagnosis coding was slightly higher for surgeries occurring before 2016, although the overall trend between 2013 and 2018 was minimal (Figure 1).

**Figure 1.**
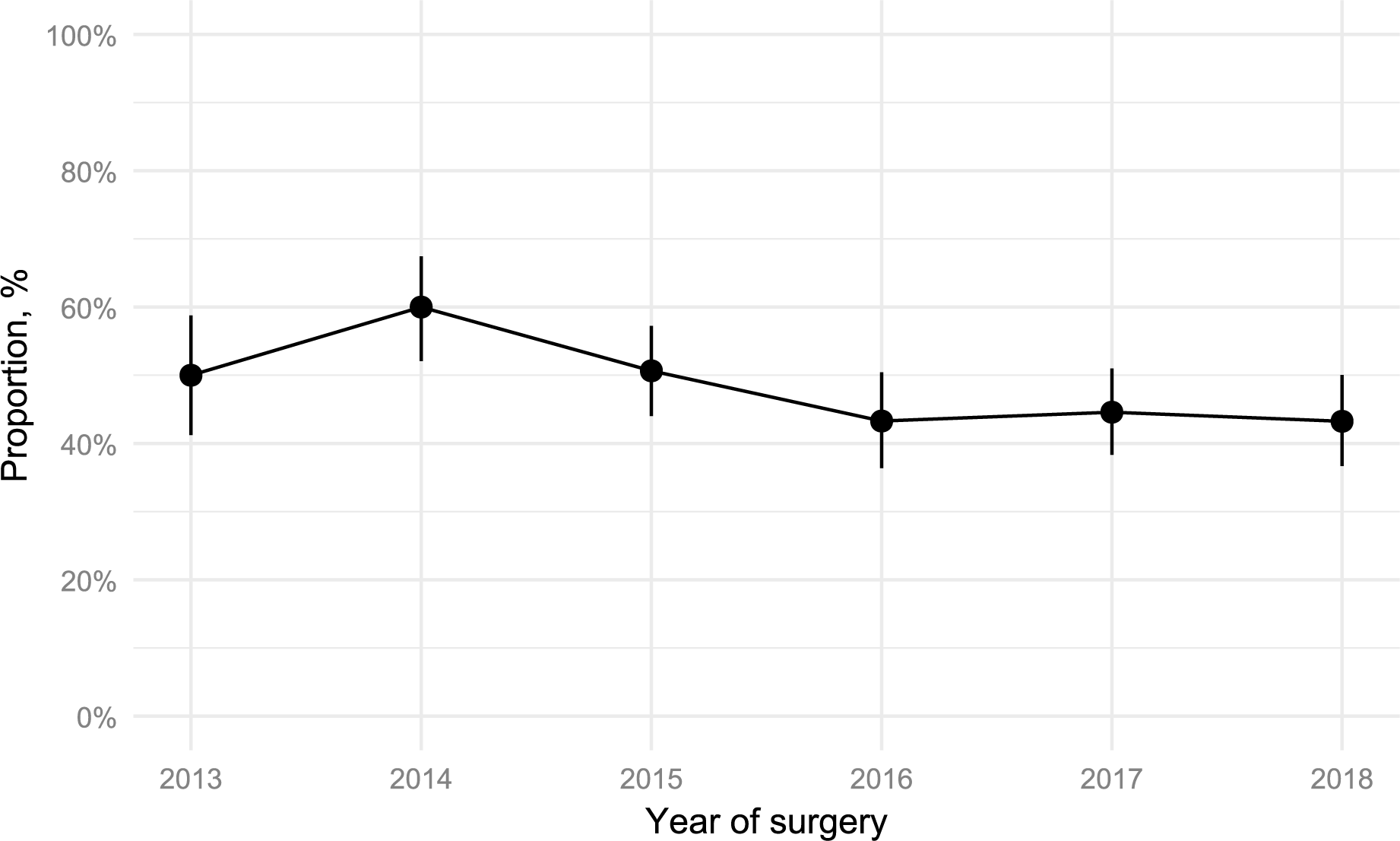
Proportion with prior OA diagnosis in primary care among patients with primary hip or knee replacement surgery. Error bars show 95% confidence intervals.

There was little difference in the rates of under-recording of OA diagnoses between hip and knee OA or between subgroups based on any observed patient characteristics. However, there was considerably more variation between GP practices (Figure 2 and Table A1).

**Figure 2.**
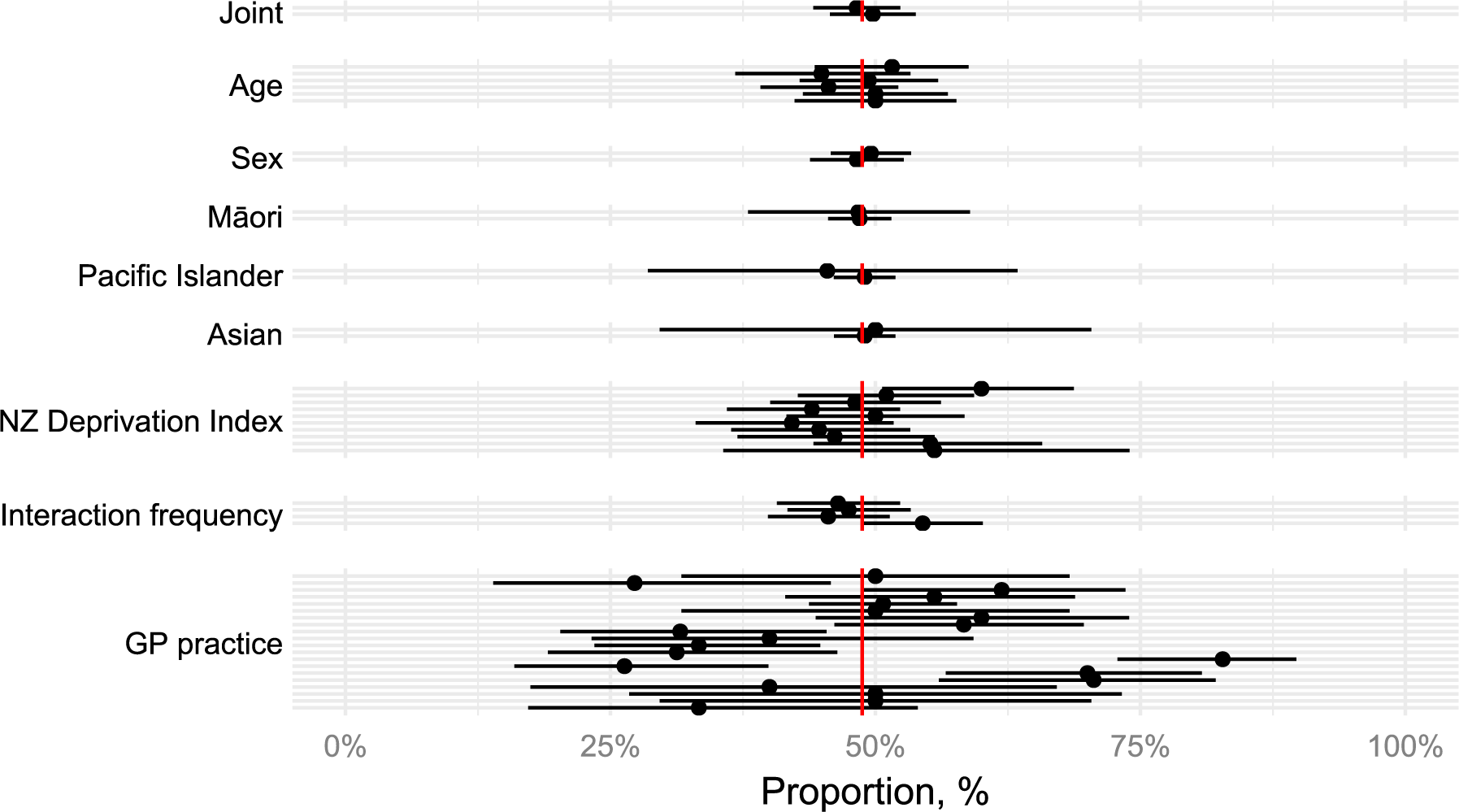
Proportion with prior OA diagnosis in primary care among patients with primary hip or knee replacement surgery, by subgroups. Vertical red line indicates the overall sample proportion. Tabular data used to create the figure are available in Table A1. Error bars show 95% confidence intervals. Abbreviations: GP=General practice; NZDep=New Zealand Index of Socioeconomic Deprivation.

The time of first recorded OA diagnosis of hip and knee OA, relative to the time of surgery, is shown in Figure 3. Among those with an OA diagnosis recorded prior to surgery, the first recorded diagnosis occurred less than one year before surgery in 27.7% of hip replacements and 16.8% of knee replacements and within 2 years of surgery for 41.5% of hip replacements and 29.7% of knee replacements. The first recorded diagnosis of OA in the primary care EHR occurred during the month *following* surgery for 16.8% of hip replacement surgeries and 20.2% of knee replacement surgeries.

**Figure 3.**
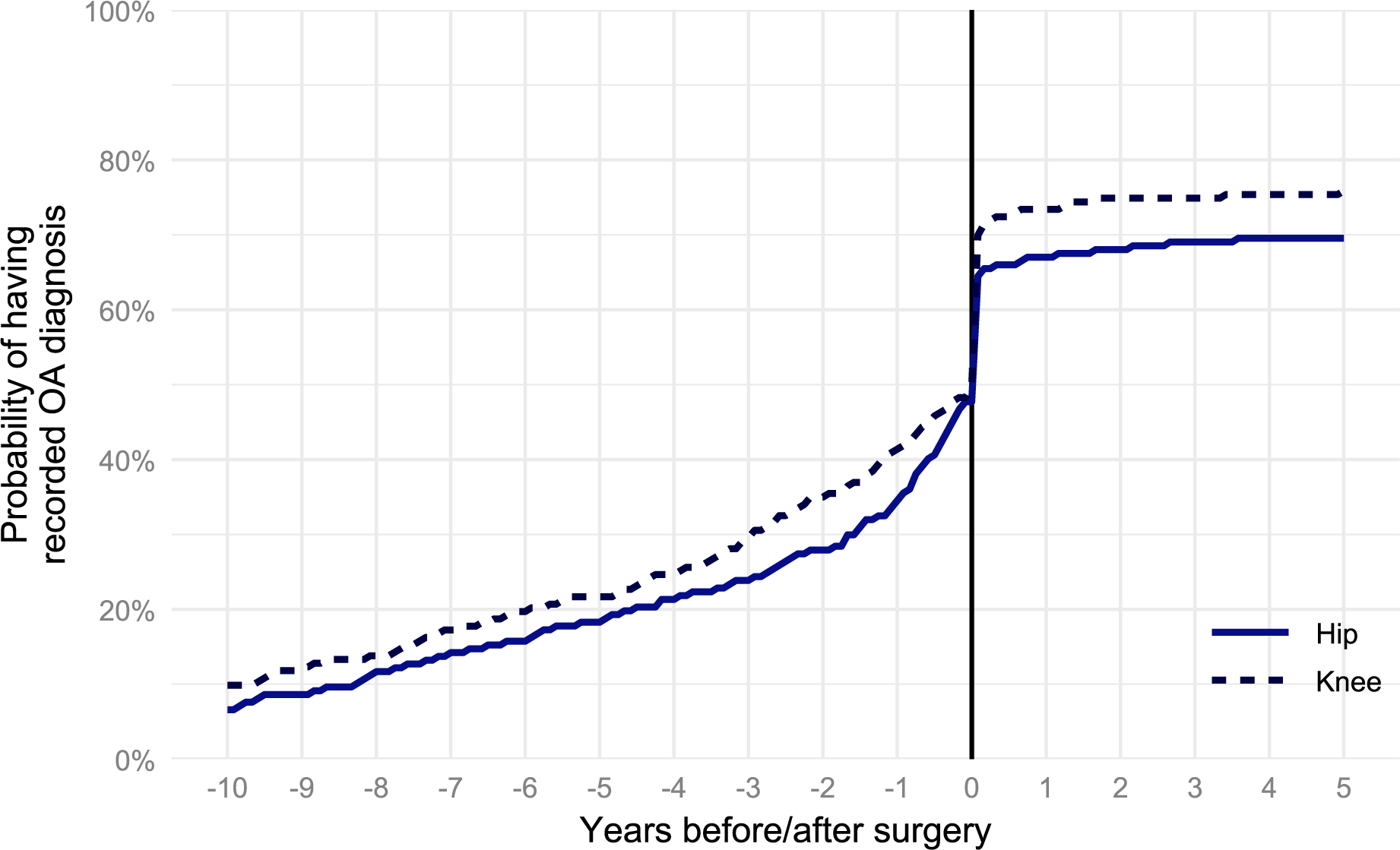
Proportion with OA diagnosis in primary care among patients with primary hip or knee replacement surgery, by time relative to surgery

## Discussion

This study investigated the extent to which OA diagnoses are under-recorded in NZ primary care, by comparison to a definitive reference standard: public hospital discharge records of patients who had total hip or knee replacement surgery for OA. We found substantial under-recording: an OA diagnosis code was missing in more than half (51.2%) of this cohort’s EHRs prior to surgery. There were no major differences observed between subgroups based on patient characteristics or OA joint, however coding behaviours varied considerably among GP practices with coding rates ranging from 26% to as high as 83% in some practices.

Strengths of this study include the use of a gold-standard reference population of patients having undergone TJR for a primary diagnosis of OA, which minimizes the risk of false positive (i.e., non-OA) cases in the reference population. Reference population challenges have been reported as a limitation within past research investigating under-recording^11^. Furthermore, by requiring all patients in our cohort to have been enrolled with the same PHO for at least 10 years prior to TJR (and including historical primary care diagnosis information for these patients as well), we ensured that any coded OA diagnoses would be captured within our data. The primary limitation of our study is that the reference population identifies only end-stage OA requiring and receiving TJR; generalizability of these results to early- and mid-stage OA is speculative. Further, the study relied on data from a single PHO covering one region of NZ, which resulted in a smaller sample than previous international studies and may limit the generalizability of our results.

Previous studies in other countries have used differing methods and reference populations, but have found similar levels of under-recording (i.e., around 50%) of OA diagnoses^9,10,15^. The methodology of the present study was most comparable with that of Yu et al.^11^. That study also used TJR as a reference population, and investigated the coding gap within EHR data retrieved from a large tranche of GP practices representing approximately 7% of the United Kingdom’s population. The coding gaps reported ranged from 47% (hip) and 56% (knee) for any OA diagnosis, to as high as 72% (hip) and 75% (knee) for joint-specific OA coding.

A novel contribution of this study was the analysis of the time between the first recorded coding of OA diagnosis and the provision of TJR. OA is a chronic condition that is ideally managed in primary care for some years before eventual need, in some cases, for TJR^4,5^. Despite this, we found that the first recorded OA diagnosis code occurred within the 2 years prior to TJR in around one-in-three cases. Orthopaedic referral within the public healthcare system in NZ is aided by prior OA diagnosis, which may explain this higher rate of OA coding shortly before TJR. In line with previous studies showing a discrepancy between narrative and coded OA diagnoses^9,10^, it is likely that in some cases OA diagnoses may have been made earlier, but not coded in EHRs at the time of diagnosis. Unlike in secondary care, primary care remuneration does not depend on accurate coding of OA diagnoses, so there is little incentive for coding unless considered necessary for effective case management.

Some GPs have reported assigning lower priority to OA, especially in situations where patients present with multiple medical needs or when faced with multi-level challenges such as time pressure and system barriers^8,20,21^. A self-reported lack of confidence in making a clinical diagnosis and a reliance on imaging to confirm diagnosis are other factors that may also impede GPs’ ability to make an accurate early OA diagnosis^8,21,22^. In other cases, GPs have described playing down or avoiding making an OA diagnosis in primary care due to the negative connotations associated with the OA label, often in an attempt to manage patient fear and disappointment^8^. Given long wait times for surgery and a lack of funded non-operative care, GPs may see little benefit in giving patients a formal OA diagnosis. Taken together, these factors may influence GPs’ decisions to defer to radiologists and orthopaedic surgeons for definitive diagnosis^22^, which may have contributed towards the observed coding behaviours; in particular, the share of patients with OA diagnosis recorded only *after* TJR. Further research is warranted to explore what may drive under-coding of OA in the NZ and international primary care context.

Care planning and resource allocation should be informed by accurate understanding of the prevalence, distribution, and management of health conditions. Our findings suggest OA is much more prevalent in primary care than implied by diagnostic coding alone, especially when considering that those who undergo TJR (one-sixth of the NZ population^23^) represent a minority of the hip and knee OA population, and presumably are those more likely to get diagnosed and coded. Our estimates of under-coding are therefore conservative, thus we strongly advise against relying on diagnostic coding as a basis for estimating OA burden in primary care.

Beyond recommending that prompt coding of OA diagnosis be prioritised in primary care settings, the observed differences in coding behaviours between GP practices involved within this study may provide material pathways for future research. By examining the knowledge, systems and processes that may underpin the coding behaviours of higher-performing GP practices, identification of the factors that influence under-recording may lead to strategies for redress.

## Conclusion

The findings of this study indicate that there is substantial under-recording of OA diagnoses in NZ primary care. Efforts to improve the consistency and timeliness of coding of hip and knee OA diagnoses are warranted.

## Acknowledgements

The authors would like to acknowledge the CMOR CREW (Collaborative Research Writing) process and writing retreat in the enabling the research and production of the manuscript.

## IDI Disclaimer

These results are not official statistics. They have been created for research purposes from the Integrated Data Infrastructure (IDI) which is carefully managed by Stats NZ. For more information about the IDI, please visit https://www.stats.govt.nz/integrated-data/.

## Contributions

**Ross Wilson:** Writing – review & editing, Writing – original draft, Methodology, Formal analysis, Conceptualization, Funding acquisition, Visualization.

**Rory McMahon Christopherson:** Writing – review & editing, Writing – original draft, Conceptualization.

**Yana Pryymachenko:** Conceptualization, Methodology, Writing – review & editing, Writing – original draft.

**Richelle Caya:** Writing – review & editing, Conceptualization.

**Lívia Gaspar Fernandes:** Writing – review & editing, Conceptualization.

**Yen Wei Lim:** Writing – review & editing, Conceptualization.

**Moody Gayed:** Writing – review & editing, Conceptualization.

**Andrés Pierobon:** Writing – review & editing, Conceptualization.

**Tony Dowell:** Conceptualization, Investigation, Methodology, Funding acquisition, Writing – Review and Editing.

**Ben Darlow:** Conceptualization, Investigation, Methodology, Funding acquisition, Writing – Review and Editing.

**Jayden MacRae:** Conceptualisation, Investigation, Methodology, Funding acquisition, Writing – Review and Editing, Data Curation.

**J. Haxby Abbott:** Writing – review & editing, Supervision, Project administration, Methodology, Conceptualization, Investigation, Funding acquisition.

## Appendix A Supplementary Material

**Table A1:**
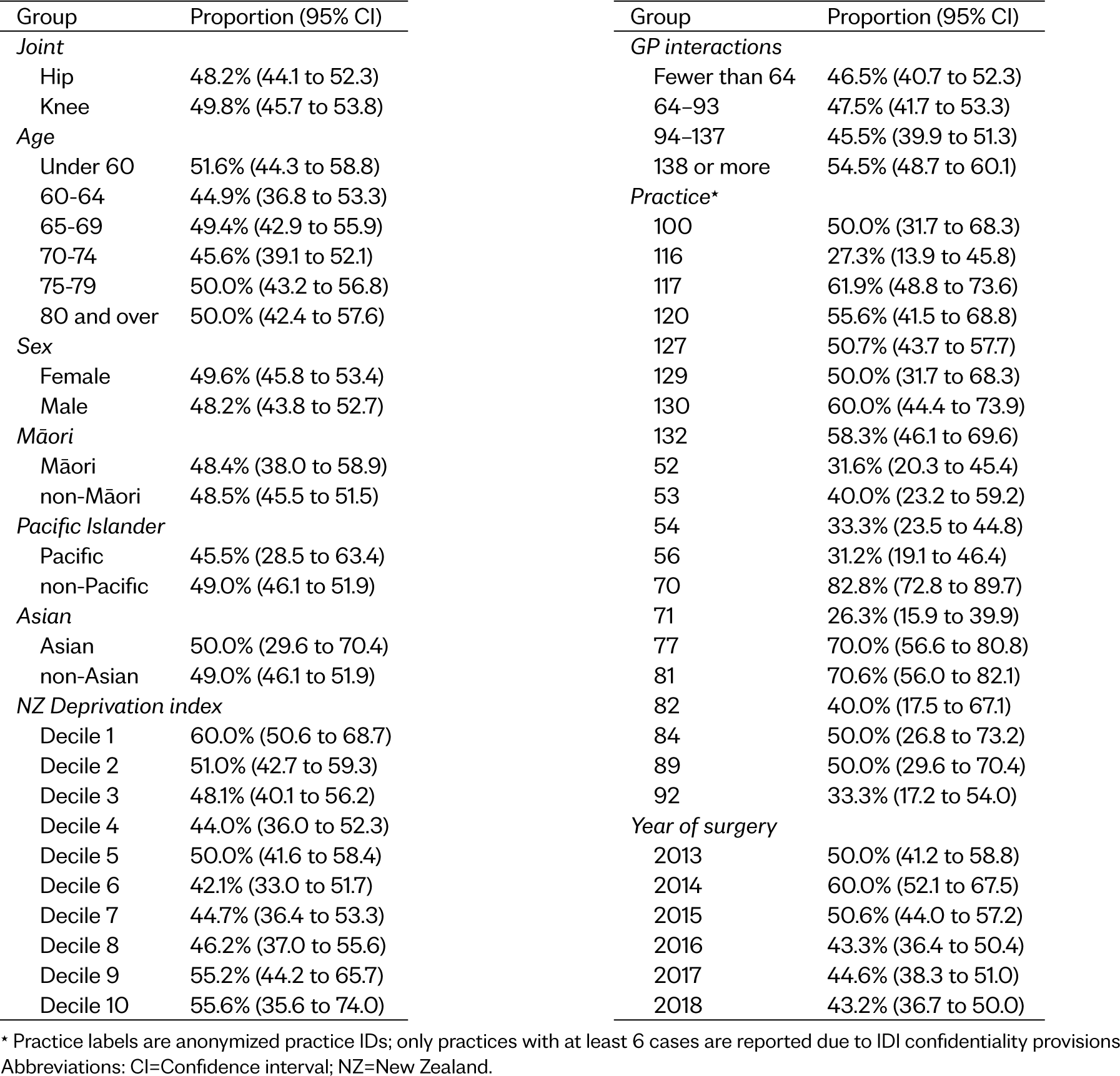
Proportion with prior osteoarthritis diagnosis in primary care among patients with primary hip or knee replacement surgery, by subgroups.

